# Cardiometabolic health and the timing of habitual exercise in the All of Us Research Program

**DOI:** 10.64898/2026.03.16.26348509

**Authors:** Prem Patel, Connor Riegal, Asanish Kalyanasundaram, Manjot Singh, Keerthenan Raveendra, Michael Y. Mi, Usman A. Tahir, Jeremy M. Robbins, Gaurav Tiwari, Nicholas S Peters, Tamar Sofer, Daniel B. Kramer, Robert E. Gerszten, Prashant Rao

## Abstract

While the cardiometabolic benefits of exercise volume and intensity are well established, the clinical significance of exercise timing remains poorly understood, largely due to the limitations of short-term accelerometry. We leveraged minute-level heart rate data from 14,489 participants from the *All of Us Research Program* to define habitual exercise timing over a one-year period. Compared to daytime exercise, habitual morning exercise was associated with lower odds of coronary artery disease (OR 0.69; CI 0.55–0.87), hypertension (OR 0.82; CI 0.72–0.94), type 2 diabetes (OR 0.70; CI 0.58–0.85), hyperlipidemia (OR 0.79; CI 0.69–0.90), and obesity (OR 0.65; CI 0.55–0.77). These associations were independent of total physical activity volume and remained consistent across hour-of-day analyses, with the lowest risk nadir occurring between 07:00-08:00 for coronary artery disease. These findings suggest that exercise timing may represent a distinct, underappreciated dimension of exercise behavior linked to cardiometabolic health.

## Introduction

Regular physical activity and exercise are key non-pharmacologic strategies for the prevention of cardiometabolic disease.^1–4^ While existing clinical guidelines prioritize exercise volume and intensity,^5,6^ emerging evidence that metabolic and cardiovascular processes are under circadian regulation suggests that exercise timing may represent an important, yet underutilized, dimension of exercise behavior.^7–12^ Despite these mechanistic findings, large-scale human data remain limited. Prior population-based analyses have relied almost exclusively on self-report or short-duration accelerometry, which lacks the longitudinal resolution to capture long-term habitual patterns and fails to distinguish structured, purposeful exercise from incidental daily movement.^13–18^

The widespread adoption of commercial wearables now enables the near-continuous monitoring of physiological signals at a population scale, enabling the characterization of habitual behaviors over months or years.^19,20^ However, no large-scale study has leveraged long-term, heart rate-defined physiology to investigate the association between habitual exercise timing and cardiometabolic disease. To address this gap, we analyzed one year of continuous Fitbit data from 14,489 participants in the *All of Us Research Program* (AoURP).^21^ We characterized individual-level patterns of habitual exercise timing and evaluated their associations with major cardiometabolic diseases, including coronary artery disease, hypertension, and type 2 diabetes.

## Methods

### Study population

Participants aged ≥18 years were enrolled in the All of Us Research Program (AoURP) following informed consent at participating clinics and regional medical centers. Detailed descriptions of the AoURP design, enrollment procedures, and data harmonization have been published previously.^21^ We used data from the AoURP Curated Data Repository (CDR) release R2024Q3R5, accessed through the Researcher Workbench.

Among 53,728 participants with linked Fitbit wearable data, 48,771 also had linked electronic health record (EHR) data, defined by the presence of at least one clinical domain (condition, procedure, drug, measurement, or visit occurrence) (CONSORT Diagram, **Supplemental Figure 1**). To ensure stable estimation of habitual exercise patterns, we restricted analyses to participants with at least 30 valid days within the first six months of the monitoring period. A valid day was defined as ≥600 minutes of non-missing heart rate data and ≥100 recorded steps.

### Fitbit wearable data

Participants who provided consent to participate in AoURP and to share EHR data were offered the opportunity to link their Fitbit account through the Bring Your Own Device program.

Participants consented to share all historical and prospective Fitbit data associated with their account. Fitbit data were collected as minute-level heart rate measurements and daily step counts. All datetime fields were subjected to random date shifting between 1 and 365 days to preserve privacy.

### Physiological definition of exercise episodes

Exercise was defined using minute-level heart rate data to capture periods of sustained physiological exertion consistent with structured exercise. For each participant, an observed maximal heart rate was defined as the highest recorded 1-minute heart rate value during the 1 year monitoring period. An acute bout of exercise (or exercise episode/session) was identified using a sliding 15-minute window; an exercise episode was determined if the mean heart rate was ≥60% of the participant’s observed maximal heart rate for at least 15 consecutive minutes. Sensitivity analyses were conducted using alternative thresholds (≥50% and ≥70% of observed maximal heart rate). The primary 60% threshold was selected following visual assessment of heart rate intensity distributions (**Supplemental Figure 2**).

To reliable capture an exercise episode, we assessed concordance between heart rate-derived exercise and total physical activity (ascertained by Fitbit). Total exercise minutes were compared with total physical activity minutes over the first year of monitoring. For each participant, we calculated the log-transformed ratio of heart rate-derived exercise minutes to total physical activity minutes as a summary measure of agreement. To minimize the influence of discordant and potentially unreliable signals, analyses were restricted to the central 55% of the log-ratio distribution of these two measures resulting in a final analytic cohort of 14,489 participants (**Supplemental Figure 3**).

In the final cohort, heart rate-derived exercise minutes were moderately correlated with total physical activity minutes (Spearman r = 0.52) (**Supplemental Figure 3**). This level of correlation is expected, as sustained elevations in heart rate reflect a structured bout of acute exercise, whereas total physical activity minutes include sporadic movement accumulated throughout the day, as well as light-intensity activity that may not meet heart rate defined exercise thresholds.

### Exercise timing phenotypes and temporal gradients

We characterized exercise timing using two complementary approaches:

1) Unsupervised clustering of participants according to their exercise timing patterns: For each participant, we constructed a 1,440-element vector (60 minutes × 24 hours) quantifying the total number of exercise episodes occurring at each minute-of-day across the monitoring period. Each element therefore represented the cumulative frequency of exercise at that specific clock minute. Vectors contain the percentage of total exercise episodes that occurred within that minute per participant. Unsupervised K-means clustering with Euclidean distance was applied to identify exercise timing phenotypes. The number of clusters (K=5) was selected a priori using the elbow criterion and confirmed by silhouette width (**Supplemental Figure 4**). Cluster stability was evaluated by comparison with a K=3 solution, which yielded highly similar patterns (**Supplemental Figure 5**). Based on overlap in timing profiles and participant characteristics, clusters were collapsed into three final phenotypes: Morning, Daytime, and Evening exercise timing groups.
2) Hour-of-day modeling: Exercise timing was also examined independently of clustering using an hour-of-day approach. The same 14,489 participants that were included in the cluster analysis were used in this hour-of-day modeling. For each participant and round hour, e.g., 5:00-5:59, 6:00-6:59, etc., we calculated the percentage of total exercise episodes starting within the hour and used this as the hour specific exposure variable. Associations between these hour-level timing exposures and cardiometabolic outcomes were evaluated to assess continuous temporal gradients.

### Outcomes

Primary outcomes were the prevalence of hypertension, type 2 diabetes, hyperlipidemia, obesity, coronary artery disease, and atrial fibrillation. Outcomes were defined using ICD-10 diagnosis codes and EHR problem-list entries harmonized within the AoURP common data model (**Supplemental Table 1**).

### Statistical analysis

Baseline characteristics were summarized across exercise timing phenotypes using means with standard deviations, medians with interquartile ranges, or counts with percentages, as appropriate. Group differences were assessed using analysis of variance, Kruskal–Wallis tests, or χ² tests. Associations between exercise timing clusters and cardiometabolic outcomes were examined using multivariable logistic regression models, with the Daytime exercise phenotype serving as the reference group. Models were adjusted for age and sex (Model 1), additionally income category, alcohol use, and smoking (Model 2), and total physical activity and sleep duration (Model 3). Covariate data were obtained from AoURP surveys, EHR records, and Fitbit summary measures. Odds ratios and 95% confidence intervals were reported. In secondary analyses, exercise timing was modeled at the hour level using percentage of total exercise episodes starting within the hour ([number of exercise episodes at that hour]/[total number of exercise episodes] x 100). For each participant, we calculated the percentage of total exercise episodes within each hour across the monitoring period and fit 24 separate multivariable models relating hour-specific timing to outcomes, adjusted for Model 3 covariates. Incident disease analyses were conducted using Cox proportional hazards models adjusted for Model 3 covariates and restricted to participants free of the outcome at the start of monitoring.

All statistical tests were two-sided, and significance was defined as P < 0.05.

## Data availability

All analyses were conducted in R version 4.3.2. The AoURP Resource Access Board approved this study, and all data were de-identified in accordance with AoURP privacy policies, including date shifting. De-identified data used in this study are available to approved researchers through the AoURP Researcher Workbench (https://workbench.researchallofus.org), subject to AoURP data access policies.

## Results

### Participant characteristics and longitudinal device characteristics

We analyzed minute-level heart rate data from 14,489 participants who met the criteria for adequate, long-term physiological monitoring. To ensure accurate estimation of habitual exercise patterns, we required participants to have at least 30 valid days over the course of the year, where a valid day was defined as 10 hours of non-missing heart rate data and at least 100 steps in that day.^20^ The final cohort (mean age 53.8 years; 61% women) exhibited high device adherence (median valid monitoring days: 138 days (IQR 74-243)), with near-continuous wear time on valid days (median 23.3 h/day (IQR 21.7-23.7)). Participants included in the final analytic cohort demonstrated higher baseline physical activity levels compared to those who did not meet the monitoring threshold (**Table 1**).

**Table 1.**
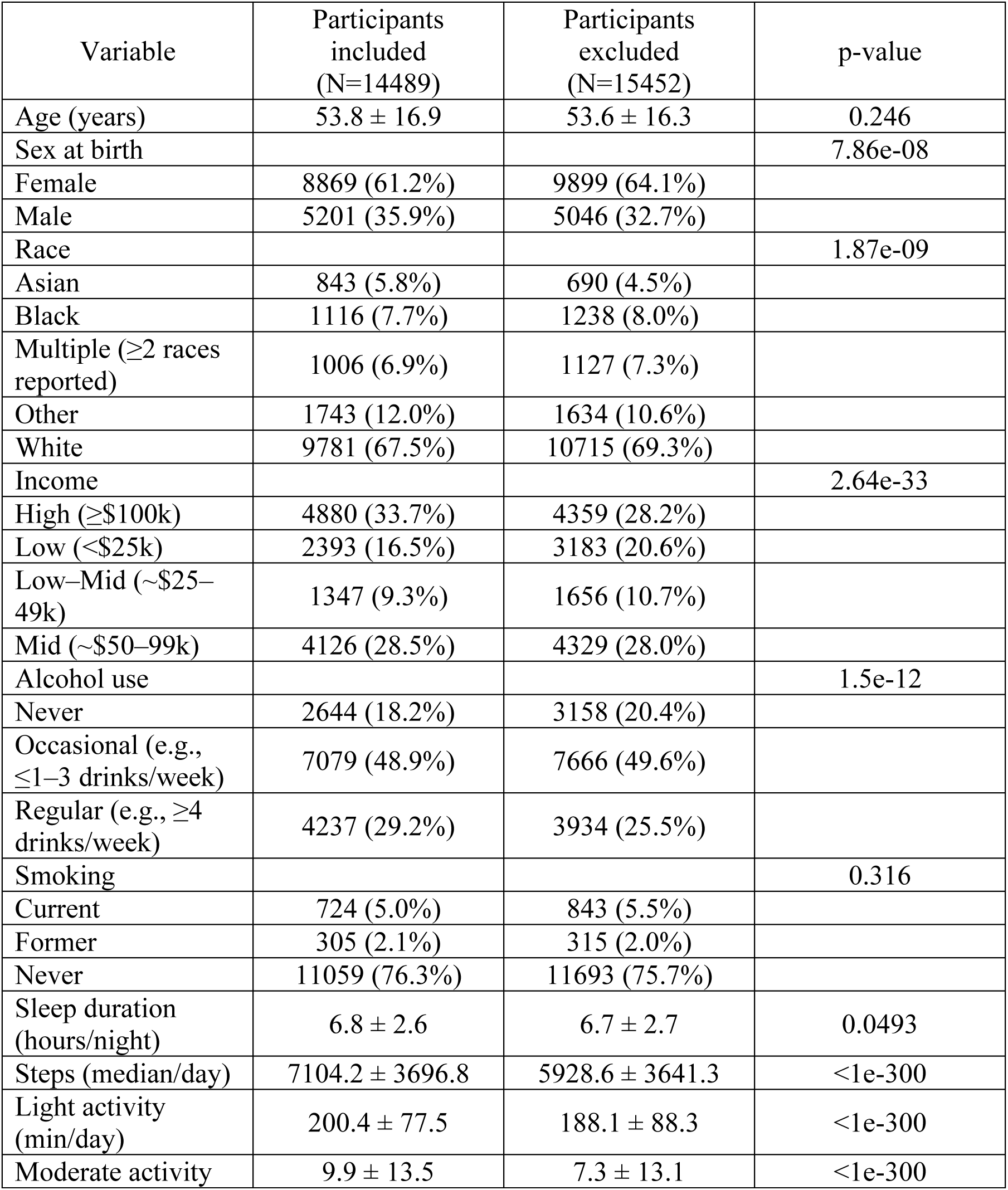
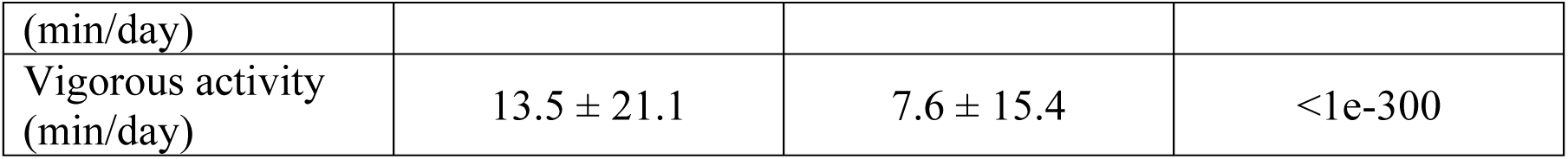
Baseline characteristics of participants with Fitbit and EHR data included versus excluded from the analytical cohort. Continuous variables are presented as mean ± standard deviation and compared using t-tests.

### Phenotypic characterization of habitual exercise timing

Using unsupervised K-means clustering of 1,440-element minute-level heart rate vectors, we identified three distinct exercise timing phenotypes: Morning (n=2,757), Daytime (n=5,728), and Evening (n=6,004) (**Figure 1**). As expected, the average time of day for exercise differed across groups (Morning (SD): 09:52 (2h:6min), Daytime: 13:30 (1h:16min), Evening: 16:15 (1h:56min)), confirming separation of timing behaviors (**Supplemental Figure 5**). Individuals in the Morning group were older, reported higher income, and achieved higher volumes of vigorous physical activity compared to the Evening group (**Table 2**).

**Figure 1.**
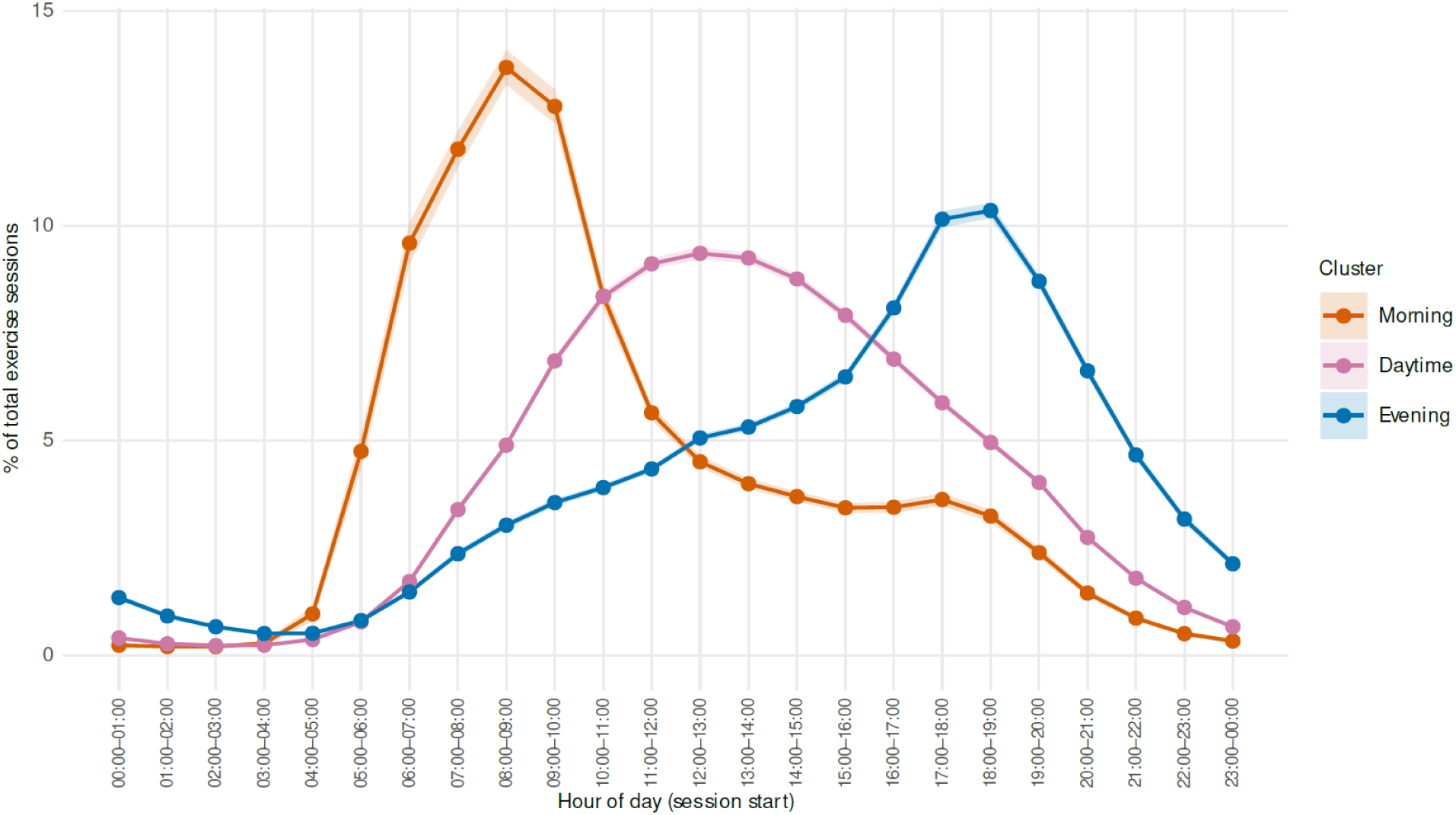
Habitual Exercise Timing Patterns Percentage of total exercise episodes occurring within each hour of the day, stratified by exercise timing clusters (Morning, Daytime, Evening). Morning group demonstrates a sharp peak in early morning hours, Daytime group shows a broad mid-day distribution, and Evening group exhibits a delayed peak in late afternoon to early evening.

**Table 2.** Baseline Characteristics of Participants by Exercise Timing Cluster Data are shown for 14,489 individuals in the 3-cluster cohort, grouped as Evening, Morning, Daytime. Continuous variables are presented as mean ± standard deviation and compared across clusters using one-way analysis of variance (ANOVA). Categorical variables are presented as number with a percentage and compared using χ² tests of independence.

| Variable | Morning<br>(N = 2,757) | Daytime<br>(N = 5,728) | Evening<br>(N = 6,004) | p-value |
| --- | --- | --- | --- | --- |
| Age (years) | 59.0 ± 15.8 | 57.0 ± 16.4 | 48.0 ± 16.1 | 1.39e-259 |
| Sex |  |  |  | 1.11e-09 |
| Female | 1652 (59.9%) | 3654 (60.9%) | 3563 (62.2%) |  |
| Male | 1065 (38.6%) | 2185 (36.4%) | 1951 (34.1%) |  |
| Race |  |  |  | 2.83e-43 |
| Asian | 173 (6.3%) | 245 (4.1%) | 425 (7.4%) |  |
| Black | 176 (6.4%) | 397 (6.6%) | 543 (9.5%) |  |
| Multiple ($\geq 2$ races reported) | 150 (5.4%) | 368 (6.1%) | 488 (8.5%) | |
| White | 1975 (71.6%) | 4324 (72.0%) | 3482 (60.8%) |  |
| Income |  |  |  | 8.96e-49 |
| Low (<\$25k) | 281 (10.2%) | 1055 (17.6%) | 1057 (18.5%) | |
| Low–Mid (~\$25–49k) | 209 (7.6%) | 556 (9.3%) | 582 (10.2%) | |
| Mid (~\$50–99k) | 811 (29.4%) | 1753 (29.2%) | 1562 (27.3%) | |
| High (≥\$100k) | 1201 (43.6%) | 1917 (31.9%) | 1762 (30.8%) | |
| Alcohol use |  |  |  | 2.99e-20 |
| Never | 525 (19.0%) | 1163 (19.4%) | 956 (16.7%) |  |
| Occasional (e.g., $\leq 1$–3 drinks/week) | 1199 (43.5%) | 2821 (47.0%) | 3059 (53.4%) | |
| Regular (e.g., $\geq 4$ drinks/week) | 937 (34.0%) | 1815 (30.2%) | 1485 (25.9%) | |
| Smoking |  |  |  | 3.97e-07 |
| Current | 150 (5.4%) | 334 (5.6%) | 240 (4.2%) |  |
| Former | 62 (2.2%) | 154 (2.6%) | 89 (1.6%) |  |
| Never | 2147 (77.9%) | 4534 (75.5%) | 4378 (76.4%) |  |
| Sleep duration (hours/night) | $6.8 \pm 2.5$ | $6.8 \pm 2.6$ | $6.7 \pm 2.7$ | 4.97e-02 |
| Steps (median/day) | $8773.8 \pm 3993.8$ | $6874.7 \pm 3570.9$ | $6541.1 \pm 3437.5$ | 1.05e-160 |
| Light activity (min/day) | $206.1 \pm 69.3$ | $203.5 \pm 77.9$ | $194.4 \pm 80.3$ | 1.51e-13 |
| Moderate activity (min/day) | $12.1 \pm 13.4$ | $10.0 \pm 14.3$ | $8.7 \pm 12.5$ | 7.49e-27 |
| Vigorous activity (min/day) | $24.8 \pm 27.7$ | $11.1 \pm 19.0$ | $10.5 \pm 17.5$ | 1.87e-211 |

### Earlier habitual exercise timing is associated with lower odds of cardiometabolic disease

In multivariable models adjusted for age, sex, socioeconomic status, lifestyle factors, total physical activity volume, and sleep duration, habitual morning exercise was consistently associated with lower odds of cardiometabolic disease (**Figure 2**). Compared with the Daytime reference group, the Morning group exhibited lower odds of coronary artery disease (OR 0.69; [95% CI 0.55-0.87], type 2 diabetes (OR 0.70; [95% CI 0.58, 0.85], obesity (OR 0.65; [95% CI 0.55-0.77], hypertension (HR 0.82; [95% CI 0.72-0.94], and hyperlipidemia (OR 0.79; [95% CI 0.69-0.90]. These associations were consistent across all nested models (**Supplemental Table 2**). Conversely, the Evening group showed directionally higher odds for hypertension, diabetes, and obesity, compared with the Daytime group. No significant associations were observed between exercise timing and atrial fibrillation, nor with the falsification endpoints (myopia, benign skin conditions) (**Figure 2**, **Supplemental Figure 6**).

**Figure 2.**
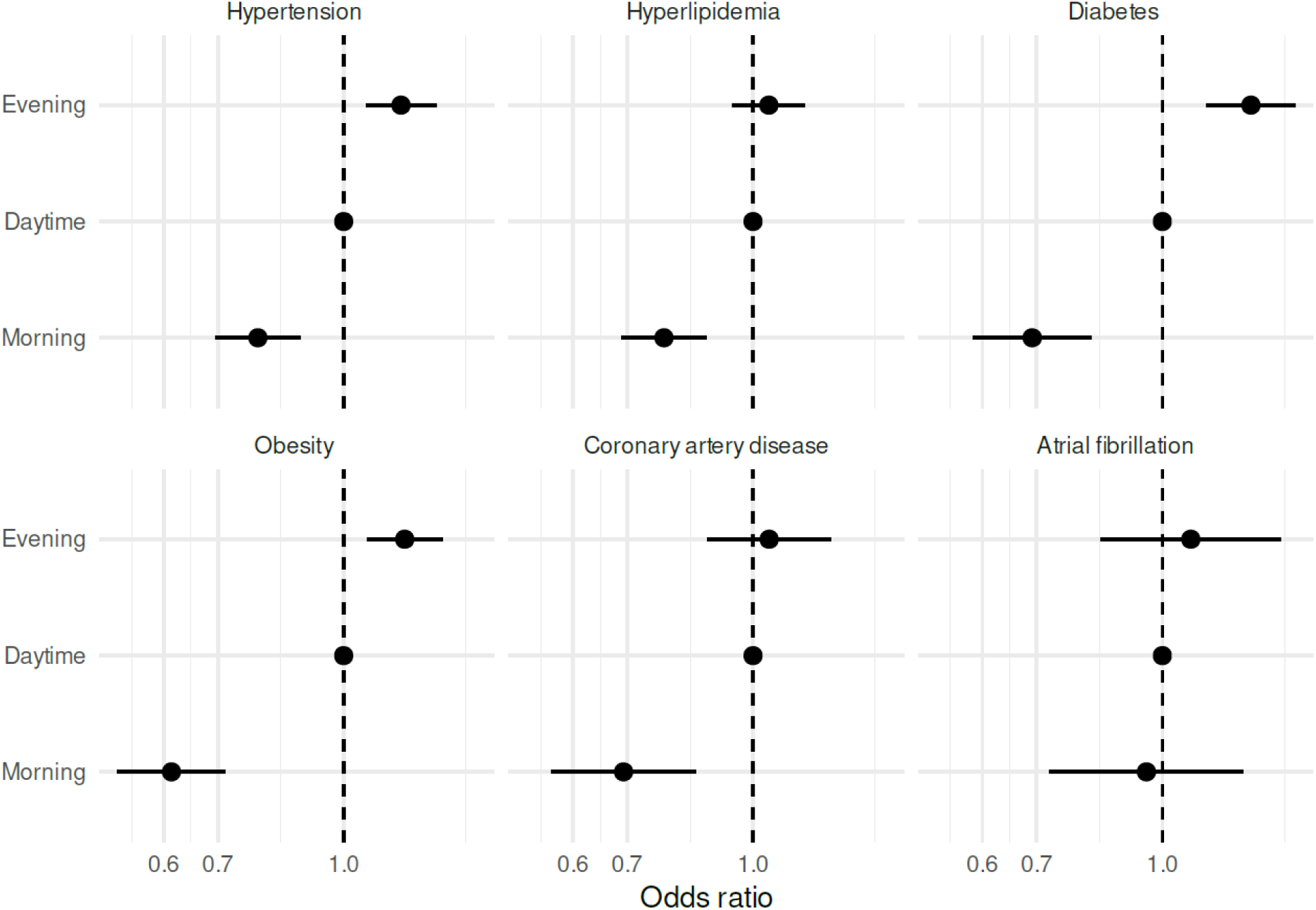
Likelihood of Conditions by Exercise Group Points indicate adjusted odds ratios (ORs) and horizontal lines indicate 95% confidence intervals from multivariable logistic regression models estimating the association of cluster membership (Evening, Morning) versus the reference group (Daytime, OR = 1) for prevalence of hypertension, hyperlipidemia, diabetes, obesity, coronary artery disease and atrial fibrillation. The x-axis is shown on a log scale, and the dashed vertical line denotes OR = 1. Models were fit and adjusted for age, sex, income category, alcohol use, smoking, sleep duration, and total physical activity.

To mitigate the potential for reverse causation, we performed longitudinal analyses restricted to incident cardiometabolic disease, excluding individuals with pre-existing conditions at the start of monitoring. In Cox proportional hazards models, the Morning group showed hazard estimates for incident type 2 diabetes, obesity, and hypertension in the same direction and magnitude as the prevalent-disease analyses, but did not achieve statistical significance given the smaller number of incident events (type 2 diabetes: HR 0.69 [95% CI 0.47-1.02], p=0.06; obesity: HR 0.71 [95% CI 0.49-1.03], p=0.07; hypertension: HR 0.77 [95% CI 0.57-1.03], p=0.08) (**Supplemental Table 3**).

### Continuous temporal gradients and sex-stratified effects

To further interrogate these associations, we next modeled exercise timing in one-hour blocks. This analysis revealed a continuous gradient across the day, with the lowest odds observed in the morning and progressively higher estimates toward the evening, consistent with the cluster-based findings (**Figure 3**). For coronary artery disease, the nadir occurred between 07:00-08:00, with ORs progressively approaching unity after ∼11:00 and exceeding 1.0 after ∼21:00. These analyses were independent of total physical activity volume, suggesting that the observed associations were not driven by differences in overall activity volume. (**Figure 3**).

**Figure 3.**
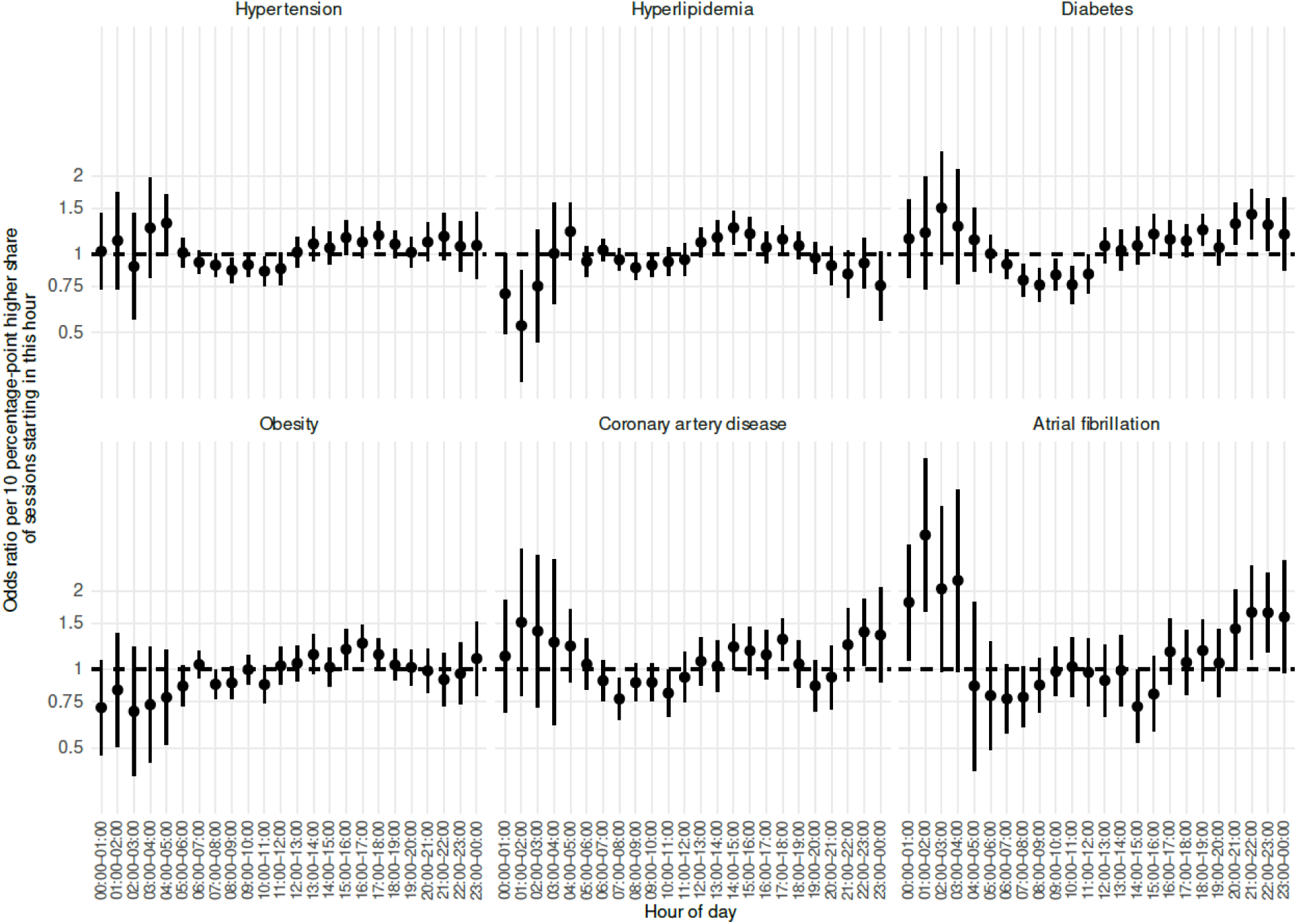
Associations of conditions with exercise by hour-of-day timing Points indicate adjusted odds ratios (ORs) and horizontal lines indicate 95% confidence intervals from multivariable logistic regression models estimating the association of exercise episode time (stratified by hour) for prevalence of hypertension, hyperlipidemia, diabetes, obesity, coronary artery disease and atrial fibrillation. ORs were estimated using multivariable logistic regression in n=14,489 participants. Models were fit and adjusted for age, sex, income category, alcohol use, smoking, sleep duration, and total physical activity.

Finally, we evaluated potential effect modification by gender. The association between earlier exercise timing and reduced cardiometabolic risk was consistent across both men and women, with no evidence of significant interaction (p for interaction >0.10 for all comparisons) (**Supplemental Table 4**).

## Discussion

In this large-scale analysis of nearly 15,000 individuals monitored over 12 months, we demonstrate a consistent association between habitual exercise timing and cardiometabolic disease. Participants who preferentially exercised in the morning exhibited lower odds of coronary artery disease, hypertension, diabetes, obesity, and hyperlipidemia. These associations were independent of total physical activity and remained consistent across both cluster-based and continuous hour-of-day analyses.

Our findings extend prior research by addressing the methodological limitations of self-report and short-duration accelerometry. In the Women’s Health Study, individuals who performed more of their total physical activity during the morning had lower odds of obesity.^22^ More recently, in the UK Biobank, participants with predominantly morning physical activity over a seven day period exhibited lower risk of coronary artery disease and stroke compared with those with midday or evening activity patterns.^15^ Similar associations have also been reported with incident dementia, although not for all-cause mortality.^17,18^ However, a major limitation of these studies centers on their reliance on short-term accelerometry which may be sensitive to transient behavioral changes and limit the ability to capture long-term habitual behavior.

Our study addresses these methodological constraints by leveraging 12 months of near-continuous heart rate monitoring, which facilitated the characterization of habitual exercise behaviors and enabled the identification of structured exercise from incidental daily movement. The convergence of our cluster-based and hour-of-day analyses, both identifying morning exercise as the period associated with the lowest cardiometabolic risk, supports the concept that exercise timing might represent a distinct and clinically relevant dimension of exercise behavior, complementary to exercise volume and intensity.

The association between morning exercise and reduced cardiometabolic burden is behaviorally and biologically plausible. Morning exercise may influence or coincide with behavioral patterns such as food timing and caloric intake, which could contribute to sustained weight management.^23,24^ In addition, morning exercise may facilitate habit formation and reduce interference from competing demands later in the day. ^25^ Beyond behavioral factors, the associations observed for morning exercise may be biologically plausible given that metabolic and cardiovascular processes exhibit diurnal variation. In small human trials, morning (08:00–09:00) aerobic exercise training elicited greater improvements in systolic blood pressure, fasting insulin, and insulin resistance than an identical afternoon (16:00–18:00) regimen, suggesting that cardiometabolic adaptations to exercise may be influenced by time of day.^26^ In mice, there is a more robust metabolic impact of exercise in the morning than at night, resulting in a higher skeletal muscle utilization of carbohydrates and ketone bodies.^10^ Morning exercise in mice also elicits greater lipolysis, increased circulating non-esterified fatty acids, and upregulation of thermogenic and mitochondrial proliferation pathways in adipose tissue.^27^

This study also highlights the utility of consumer wearables for high-resolution behavioral phenotyping at scale. By analyzing minute-level heart rate vectors over a full year, we demonstrated the feasibility of objective, longitudinal monitoring in free-living populations. However, several limitations merit consideration. The observational design precludes causal inference, although the consistency of effect sizes in the incident-disease analyses and the lack of association with negative-control outcomes mitigate concerns regarding reverse causation and residual confounding. It remains possible that these findings may reflect differences in work structure and caregiving demands. In addition, our heart rate-based definitions may not fully capture resistance exercise with prolonged rest periods, and the study population, comprised of active device users, may not generalize to populations with lower digital health literacy.

## Conclusion

Habitual morning exercise is associated with a lower likelihood of major cardiometabolic conditions, independent of total activity. These observations suggest that exercise timing may represent a previously underappreciated and modifiable dimension of cardiometabolic health. These findings motivate randomized trials to determine whether the strategic timing of exercise can be optimized to prevent and manage cardiometabolic disease.

## Source of Financial Support

M.Y.M. is supported by NIH grant K23HL171855. D.B.K reports funding from NIH and PCORI, not related to this paper, as well as consulting to Whoop, Inc. P.R is supported by NIH grant (K23HL177335-01).

## Data Availability

De identified data used in this study are available to approved researchers through the AoURP Researcher Workbench (https://workbench.researchallofus.org), subject to AoURP data access policies.

## Acknowledgements

We gratefully acknowledge *All of Us* participants for their contributions, without whom this research would not have been possible

## Supplemental Material

**Supplementary Table 1.**
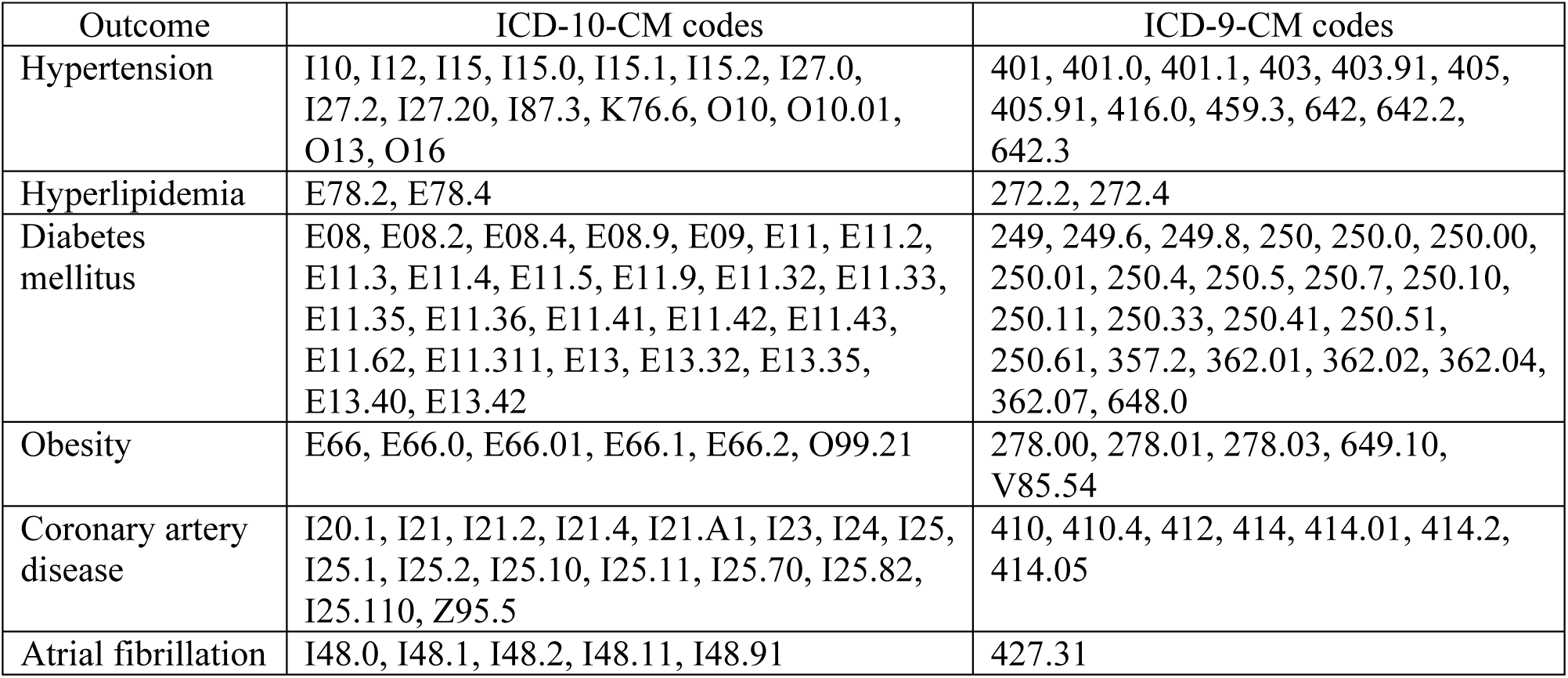
ICD-9-CM and ICD-10-CM codes used to define cardiometabolic outcomes.

| Outcome | ICD-10-CM codes | ICD-9-CM codes |
| --- | --- | --- |
| Hypertension | I10, I12, I15, I15.0, I15.1, I15.2, I27.0, I27.2, I27.20, I87.3, K76.6, O10, O10.01, O13, O16 | 401, 401.0, 401.1, 403, 403.91, 405, 405.91, 416.0, 459.3, 642, 642.2, 642.3 |
| Hyperlipidemia | E78.2, E78.4 | 272.2, 272.4 |
| Diabetes mellitus | E08, E08.2, E08.4, E08.9, E09, E11, E11.2, E11.3, E11.4, E11.5, E11.9, E11.32, E11.33, E11.35, E11.36, E11.41, E11.42, E11.43, E11.62, E11.311, E13, E13.32, E13.35, E13.40, E13.42 | 249, 249.6, 249.8, 250, 250.0, 250.00, 250.01, 250.4, 250.5, 250.7, 250.10, 250.11, 250.33, 250.41, 250.51, 250.61, 357.2, 362.01, 362.02, 362.04, 362.07, 648.0 |
| Obesity | E66, E66.0, E66.01, E66.1, E66.2, O99.21 | 278.00, 278.01, 278.03, 649.10, V85.54 |
| Coronary artery disease | I20.1, I21, I21.2, I21.4, I21.A1, I23, I24, I25, I25.1, I25.2, I25.10, I25.11, I25.70, I25.82, I25.110, Z95.5 | 410, 410.4, 412, 414, 414.01, 414.2, 414.05 |
| Atrial fibrillation | I48.0, I48.1, I48.2, I48.11, I48.91 | 427.31 |

**Supplemental Table 2:** Cluster Associations with Odds of Disease Reference group = Daytime. Values are OR (95% CI) for Morning vs Daytime and Evening vs Daytime. Model 1: adjusted for age and sex. Model 2: adjusted for age, sex, income category, alcohol category, and smoking category. Model 3: adjusted for age, sex, income category, alcohol category, smoking category, sleep duration, and total physical activity.

| Condition | Group | Cases | Model 1 OR (95% CI); P | Model 2 OR (95% CI); P | Model 3 OR (95% CI); P |
| --- | --- | --- | --- | --- | --- |
| Hypertension | Daytime | 401 | 1.00 (reference) | 1.00 (reference) | 1.00 (reference) |
| Hypertension | Morning | 396 | 0.71 (0.64–0.80); 3.44e-09 | 0.77 (0.68–0.87); 1.58e-05 | 0.82 (0.72–0.94); 3.50e-03 |
| Hypertension | Evening | 175 | 1.15 (1.05–1.27); 2.76e-03 | 1.20 (1.08–1.32); 3.74e-04 | 1.22 (1.09–1.36); 5.09e-04 |
| Hyperlipidemia | Daytime | 409 | 1.00 (reference) | 1.00 (reference) | 1.00 (reference) |
| Hyperlipidemia | Morning | 432 | 0.75 (0.67–0.84); 3.52e-07 | 0.76 (0.68–0.86); 1.08e-05 | 0.79 (0.69–0.90); 5.98e-04 |
| Hyperlipidemia | Evening | 188 | 1.04 (0.95–1.15); 3.82e-01 | 1.06 (0.96–1.17); 2.58e-01 | 1.07 (0.96–1.20); 2.27e-01 |
| Diabetes | Daytime | 182 | 1.00 (reference) | 1.00 (reference) | 1.00 (reference) |
| Diabetes | Morning | 158 | 0.62 (0.53–0.72); 1.55e-09 | 0.67 (0.57–0.79); 3.30e-06 | 0.70 (0.58–0.85); 2.25e-04 |
| Diabetes | Evening | 92 | 1.27 (1.13–1.43); 6.46e-05 | 1.32 (1.17–1.50); 1.32e-05 | 1.31 (1.14–1.51); 1.75e-04 |
| Obesity | Daytime | 106 | 1.00 (reference) | 1.00 (reference) | 1.00 (reference) |
| Obesity | Morning | 134 | 0.55 (0.47–0.63); 7.82e-17 | 0.60 (0.51–0.70); 3.25e-11 | 0.65 (0.55–0.77); 4.39e-07 |
| Obesity | Evening | 46 | 1.17 (1.06–1.30); 1.76e-03 | 1.21 (1.09–1.35); 4.36e-04 | 1.18 (1.05–1.33); 7.18e-03 |
| Coronary artery disease | Daytime | 109 | 1.00 (reference) | 1.00 (reference) | 1.00 (reference) |
| Coronary artery disease | Morning | 96 | 0.67 (0.55–0.81); 4.04e-05 | 0.69 (0.56–0.84); 3.07e-04 | 0.69 (0.55–0.87); 1.54e-03 |
| Coronary artery disease | Evening | 48 | 1.09 (0.93–1.29); 2.86e-01 | 1.06 (0.89–1.27); 4.87e-01 | 1.04 (0.86–1.27); 6.68e-01 |
| Atrial fibrillation | Daytime | 37 | 1.00 (reference) | 1.00 (reference) | 1.00 (reference) |
| Atrial fibrillation | Morning | 31 | 0.87 (0.67–1.14); 3.15e-01 | 0.95 (0.72–1.25); 7.06e-01 | 0.98 (0.71–1.34); 8.87e-01 |
| Atrial fibrillation | Evening | 14 | 1.10 (0.87–1.41); 4.25e-01 | 1.10 (0.85–1.42); 4.71e-01 | 1.17 (0.87–1.56); 2.95e-01 |

**Supplemental Table 3:** Cluster Associations with Incident Disease Reference group = Daytime. Values are OR (95% CI) for Morning vs Daytime and Evening vs Daytime. Analyses adjusted for age, sex, income category, alcohol category, smoking category, sleep duration, and total physical activity.

| Outcome | Group | N | Events | HR (95% CI) | p-value |
| --- | --- | --- | --- | --- | --- |
| Hypertension | Daytime (Ref) | 4441 | 143 | 1 | - |
|  | Morning | 2154 | 69 | 0.77 (0.57–1.03) | 0.081 |
|  | Evening | 4519 | 125 | 0.90 (0.69–1.16) | 0.406 |
| Hyperlipidemia | Daytime (Ref) | 4436 | 163 | 1 | - |
|  | Morning | 2125 | 84 | 0.89 (0.67–1.17) | 0.399 |
|  | Evening | 4633 | 137 | 0.96 (0.75–1.23) | 0.729 |
| Diabetes | Daytime (Ref) | 5265 | 97 | 1 | - |
|  | Morning | 2523 | 40 | 0.69 (0.47–1.02) | 0.063 |
|  | Evening | 5054 | 91 | 0.98 (0.72–1.34) | 0.921 |
| Obesity | Daytime (Ref) | 5011 | 106 | 1 | - |
|  | Morning | 2484 | 46 | 0.71 (0.49–1.03) | 0.074 |
|  | Evening | 4727 | 134 | 1.04 (0.79–1.37) | 0.801 |
| Coronary Artery Disease | Daytime (Ref) | 5559 | 57 | 1 | - |
|  | Morning | 2597 | 25 | 0.86 (0.53–1.38) | 0.523 |
|  | Evening | 5453 | 40 | 0.96 (0.62–1.49) | 0.866 |
| Atrial Fibrillation | Daytime (Ref) | 5814 | 28 | 1 | - |
|  | Morning | 2671 | 10 | 0.60 (0.28–1.28) | 0.188 |
|  | Evening | 5613 | 18 | 0.88 (0.46–1.69) | 0.702 |

**Supplemental Table 4:**
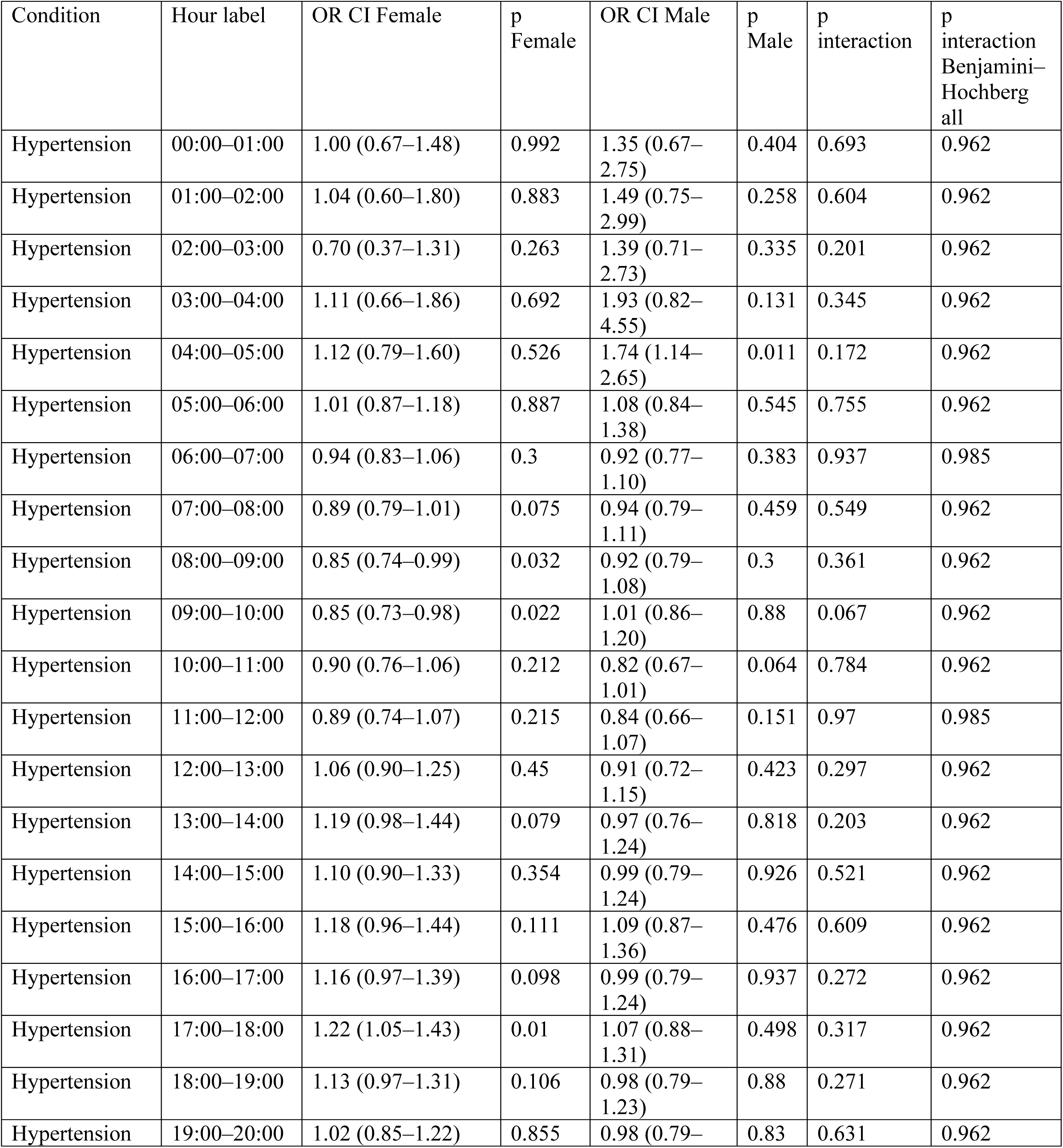
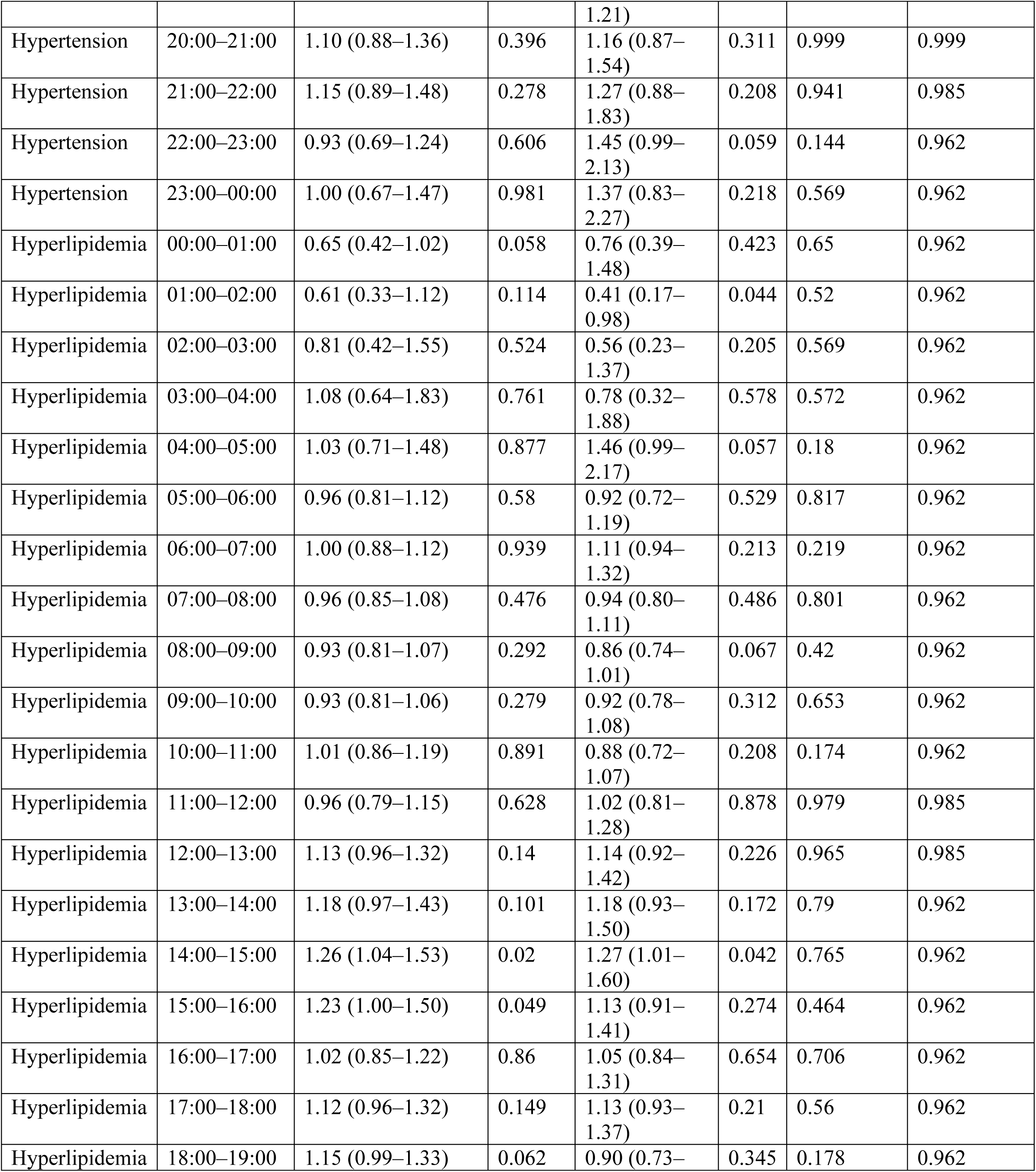
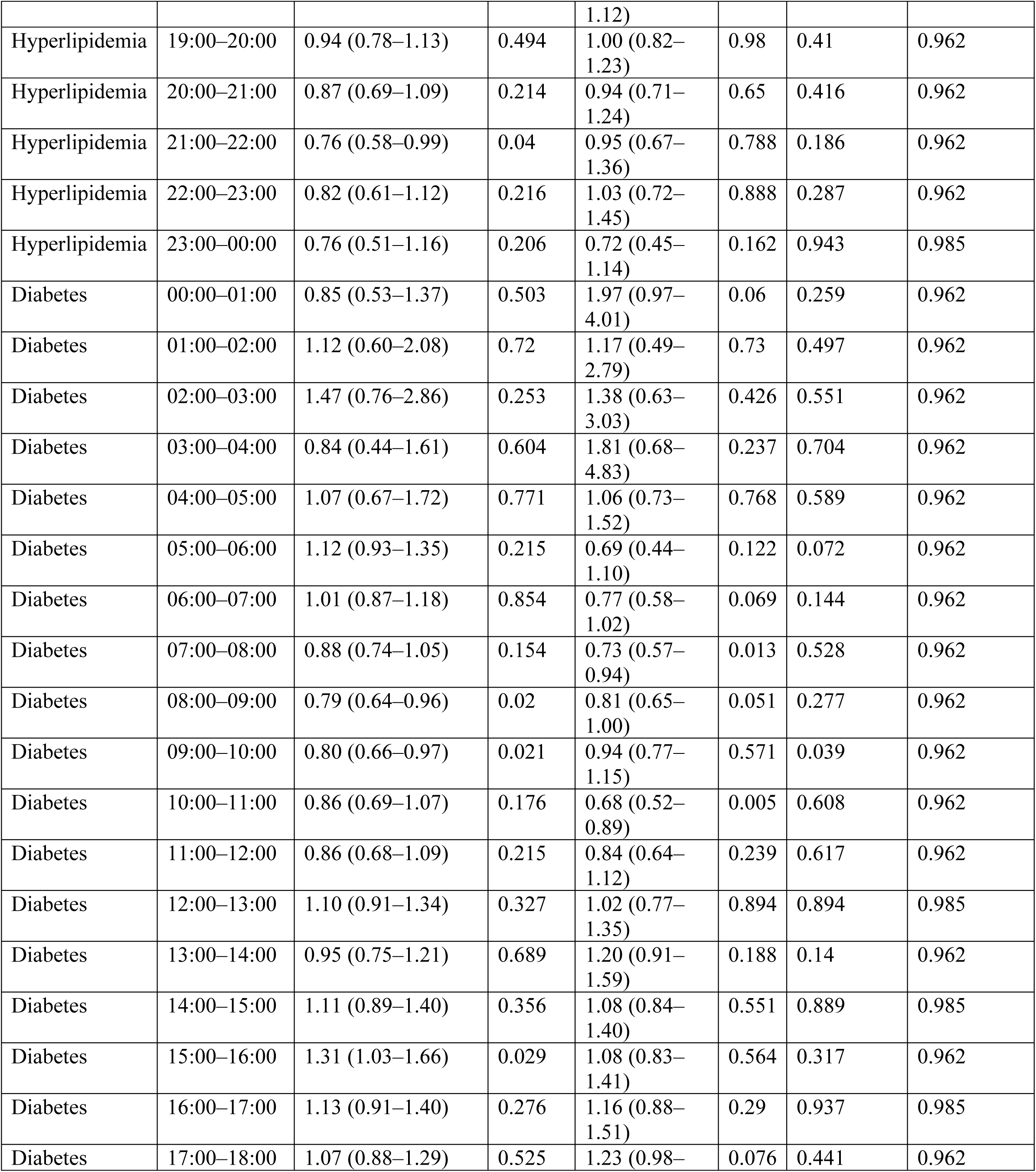
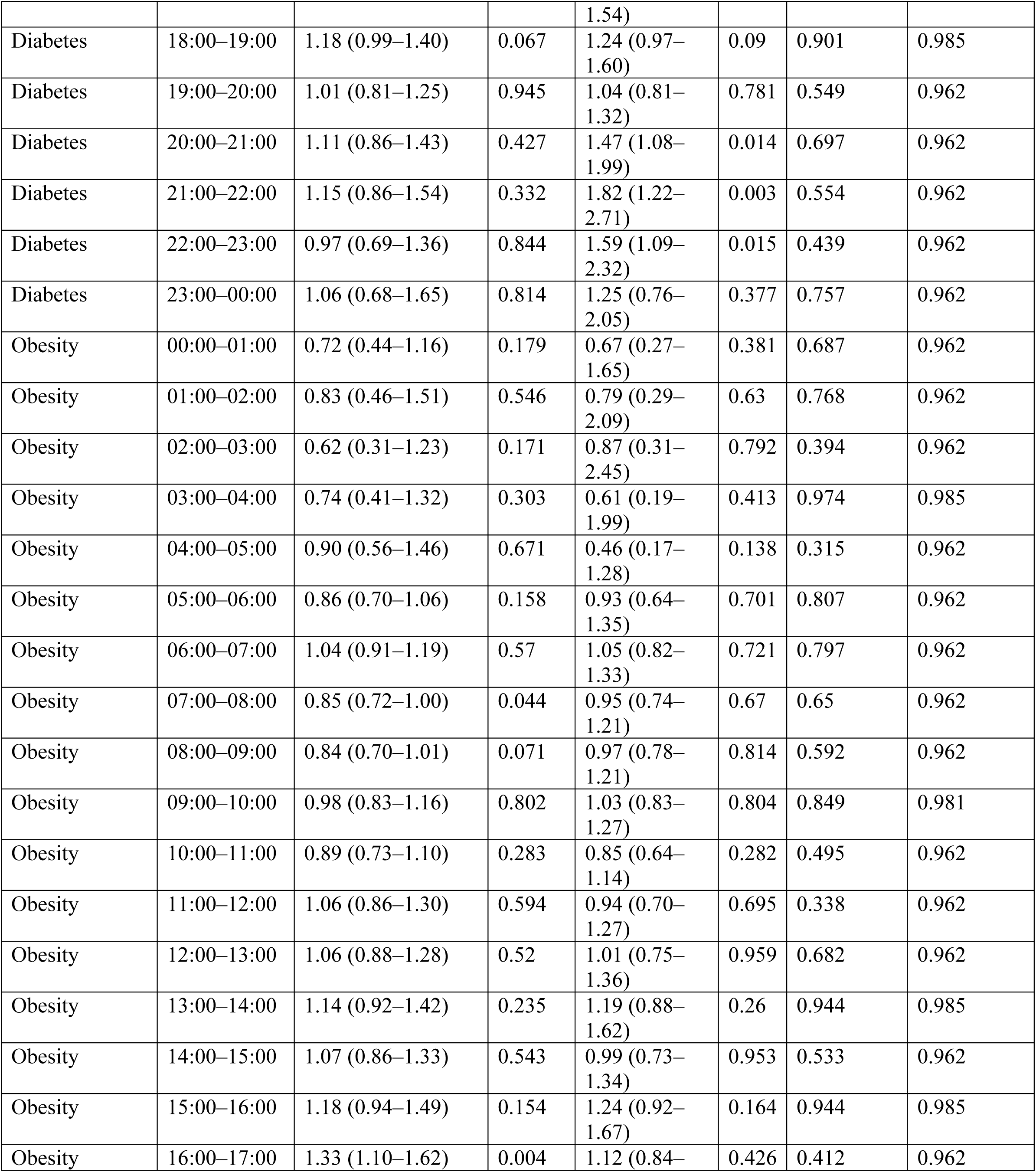
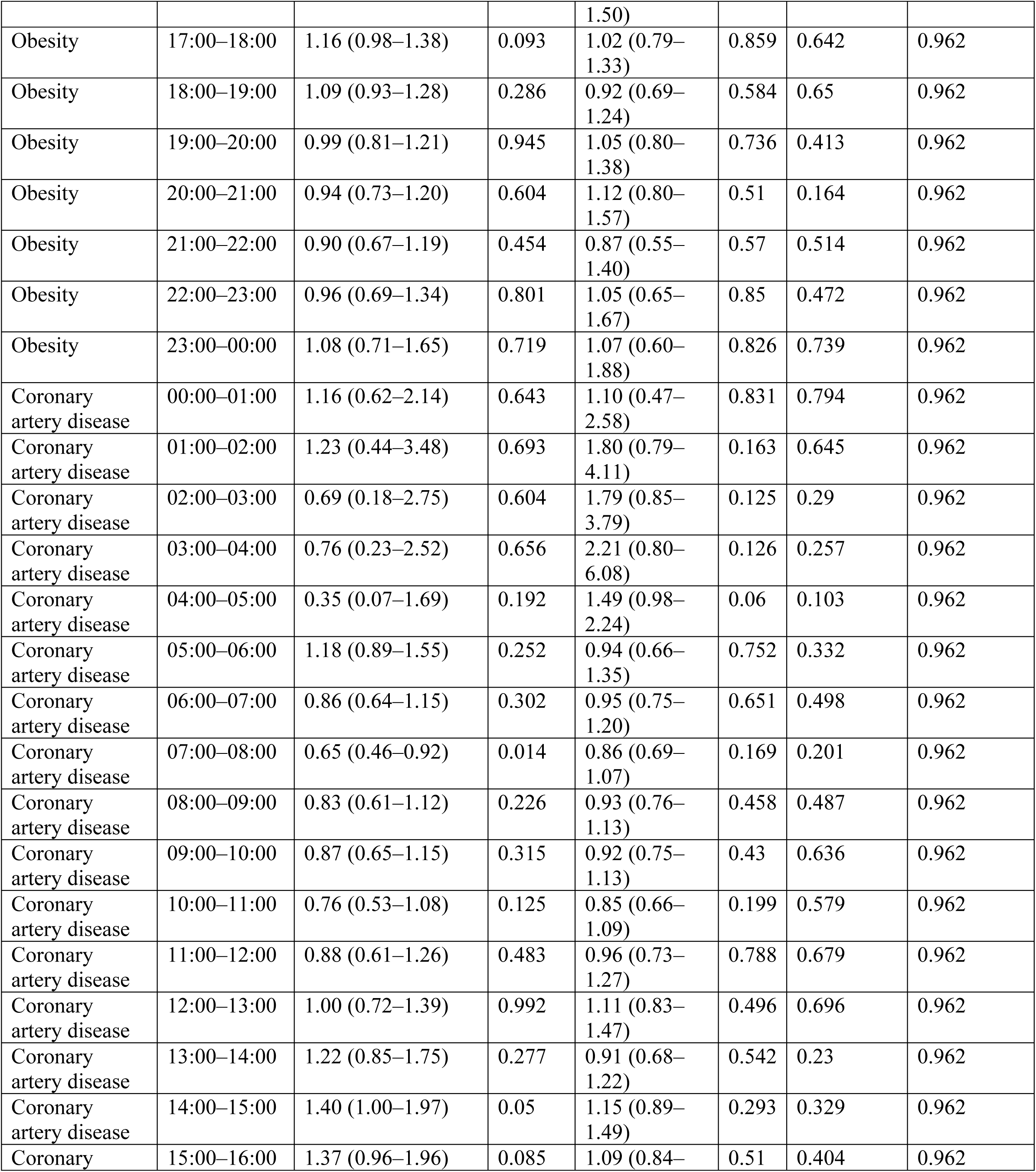
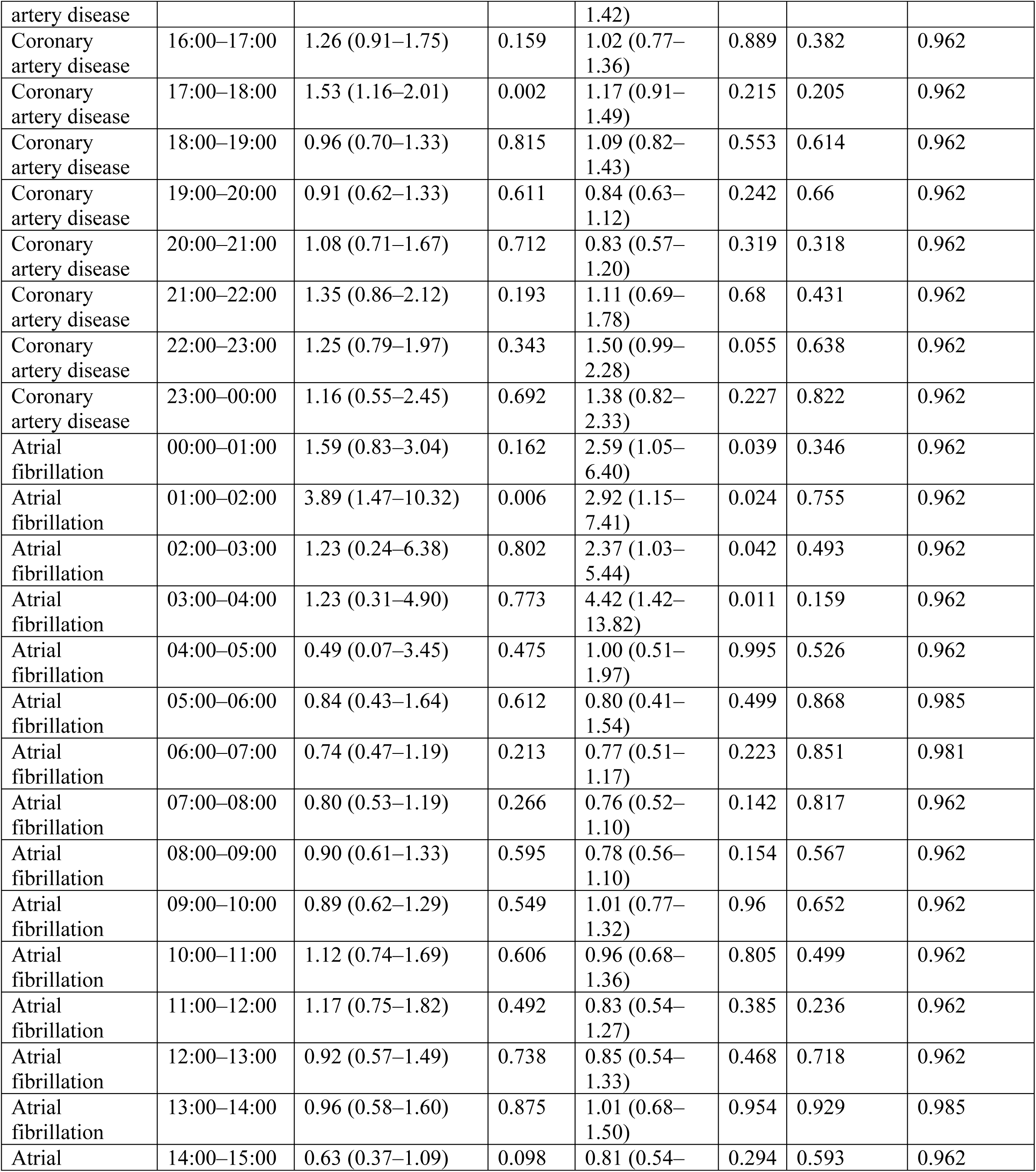
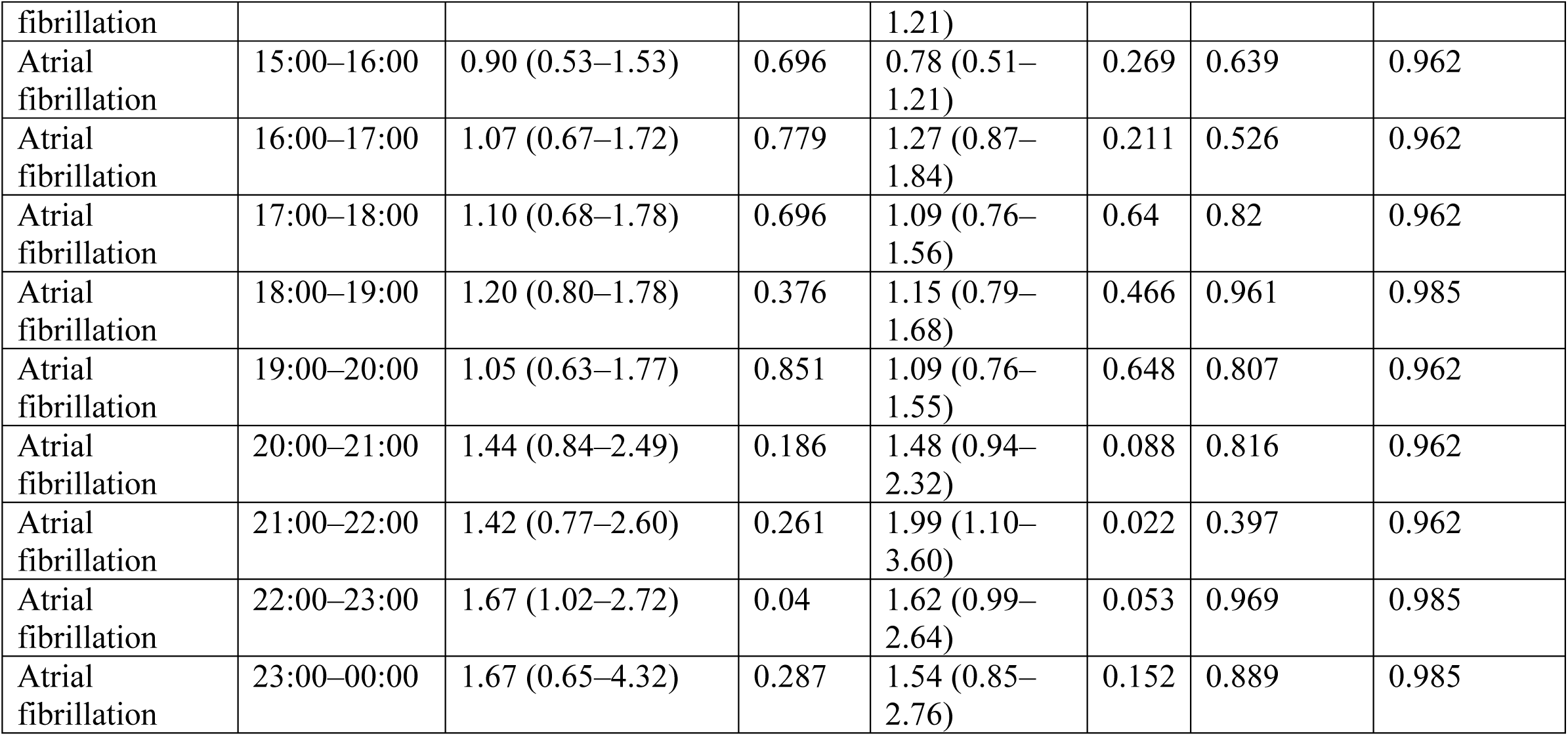
Sex-interaction and sex-stratified hour-by-hour associations.

## Supplemental Figures

**Supplemental Figure 1:**
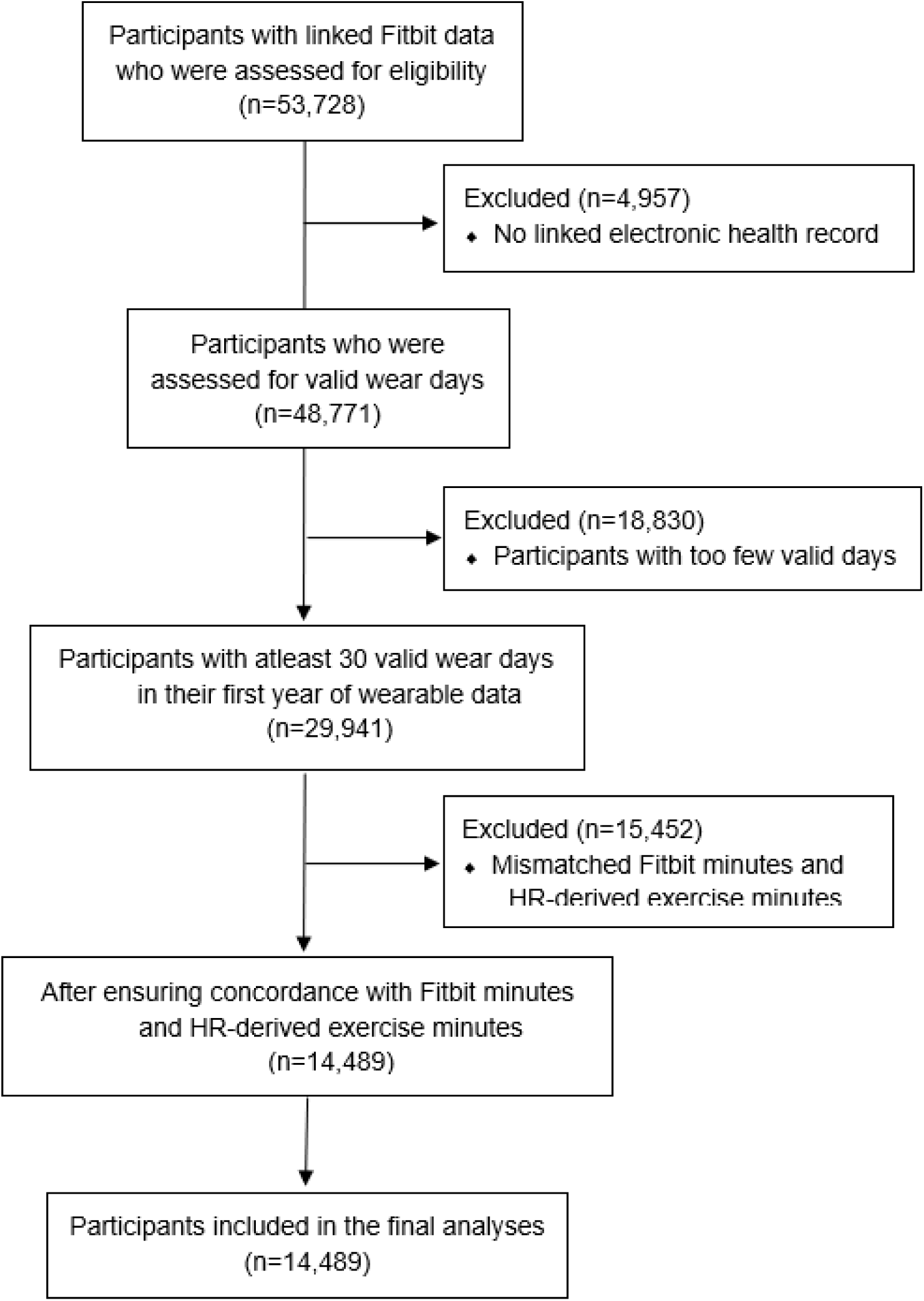
CONSORT Diagram

**Supplemental Figure 2:**
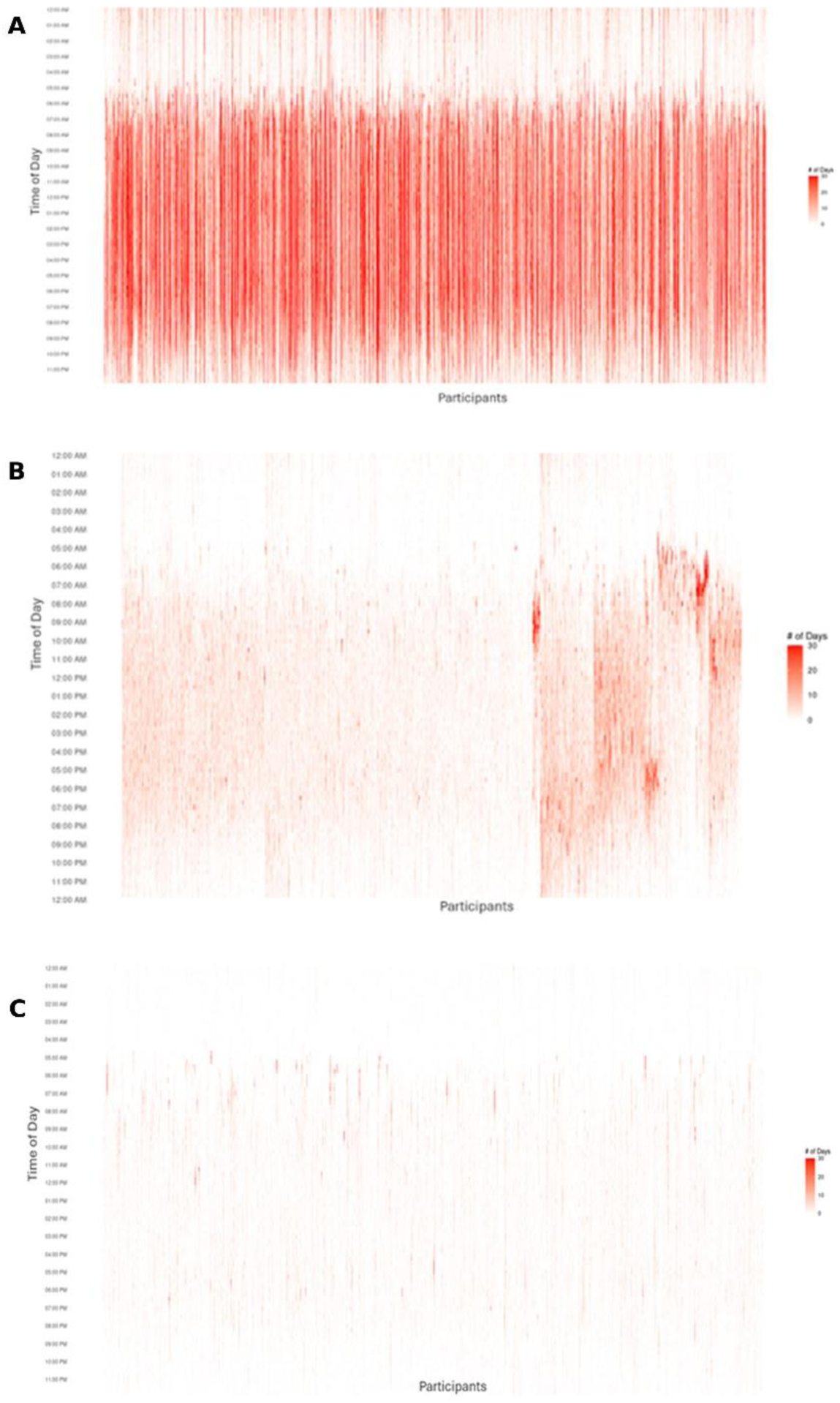
Exercise timing patterns using alternative heart rate thresholds. Heatmaps showing the distribution of exercise timing across the 24-hour day using heart rate thresholds of (A) 50%, (B) 60%, and (C) 70% of maximal heart rate. Lower thresholds (50%) resulted in denser activity patterns, whereas higher thresholds (70%) resulted in more sparse exercise.

**Supplemental Figure 3.**
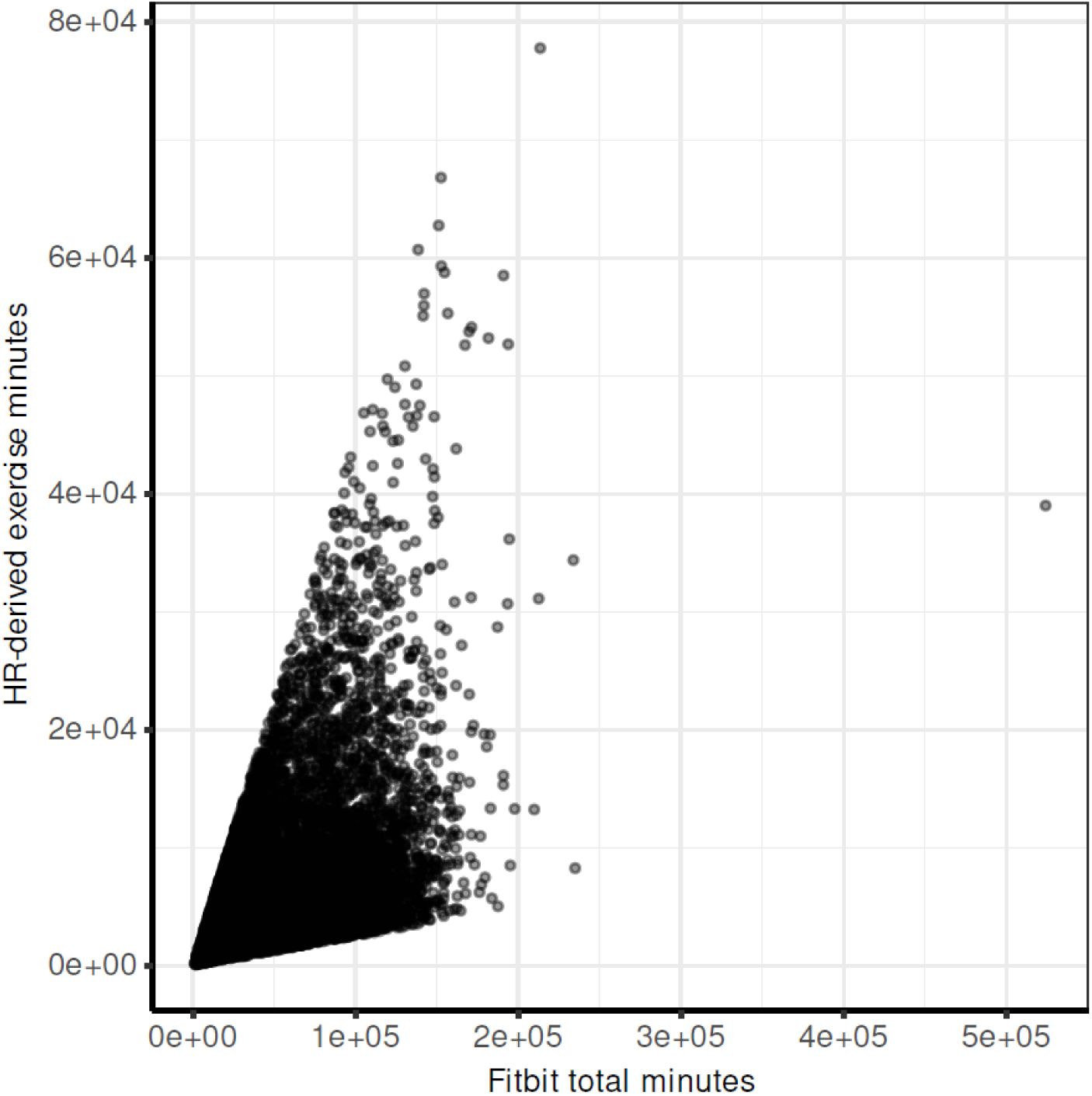
Correlation between calculated exercise minutes and total physical activity. Each point represents one participant (n=14489) and plots total HR-derived exercise minutes and total physical activity minutes over one-year. There is overall moderate agreement between Fitbit-activity and HR-based measures and was used for quality control to identify participants with discordant signals.

**Supplemental Figure 4.**
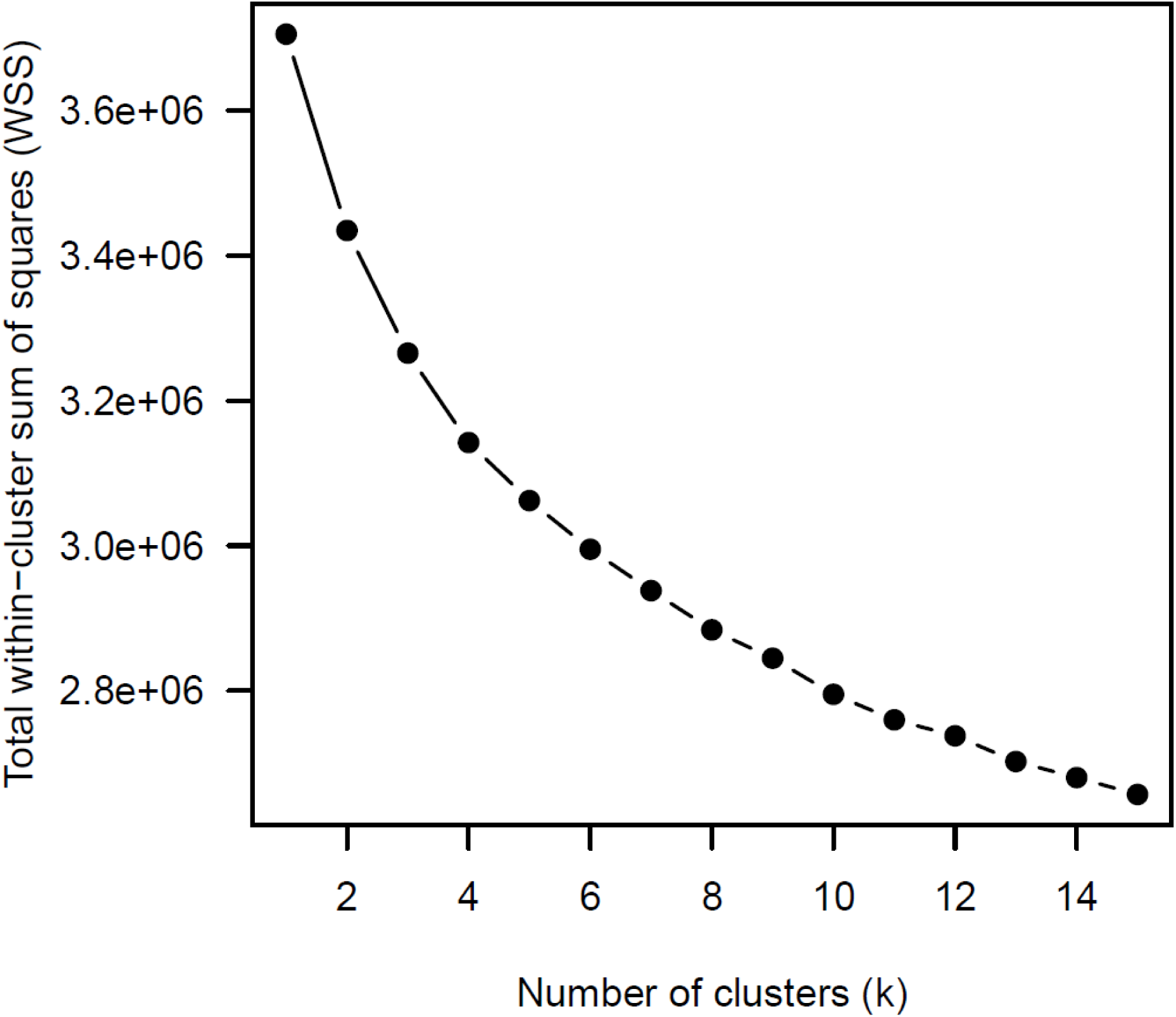
Elbow Method for Optimal Clustering Total within-cluster sum of squares is shown for k-means clustering of n=14,489 participants, evaluated across k = 1–15 clusters. Each point represents the total within-cluster dispersion of participants’ exercise-episode timing profiles. The elbow method was used to guide selection of the final k (k=5) for downstream clustering.

**Supplemental Figure 5.**
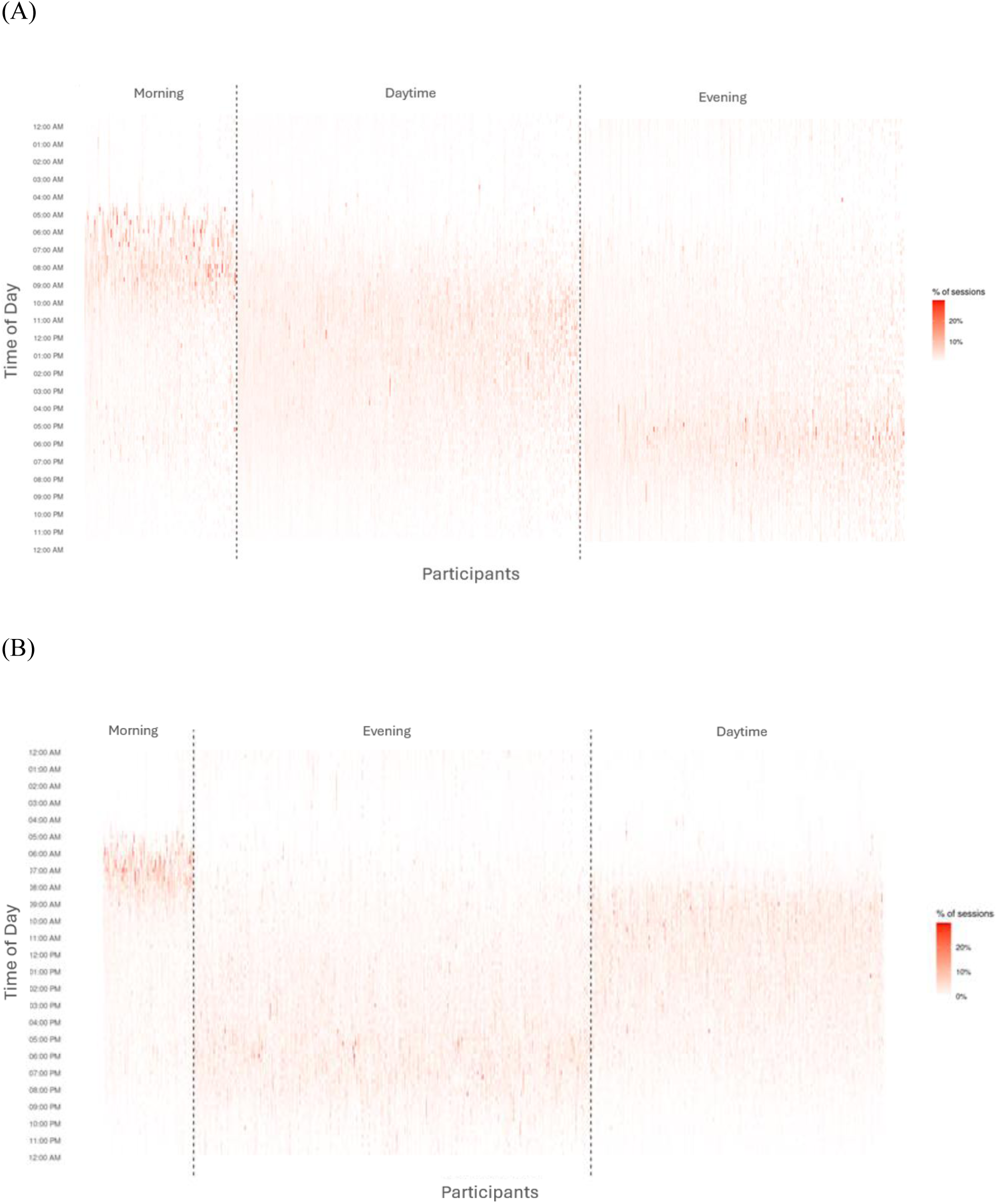
Heatmap Exercise Timing Patterns in Behaviorally Clustered Each column represents one participant’s cumulative exercise data over one-year. The heatmap shows the percentage of that participant’s detected exercise episodes occurring in each 15-min time-of-day bin. The gradient indicates the percentage of episodes (capped at a maximum of 30% of total episodes for display). (A) *K = 5 clustering with collapsed groups.* Participants were first grouped using k-means clustering (K = 5; elbow method). To facilitate interpretability and comparison with the primary analysis, adjacent clusters with similar timing distributions were combined, yielding three predominant patterns: Morning (n = 2,757), Daytime (n = 6,004), and Evening (n = 5,728). Specifically, two late-day clusters were merged to form the Evening group, and two early-day clusters were merged to form the Morning group. (B) *K = 3 clustering.* Participants were grouped directly into three clusters using k-means clustering (K = 3; elbow method), corresponding to Morning (n = 1,671), Daytime (n = 7,386), and Evening (n = 5,432).

**Supplemental Figure 6.**
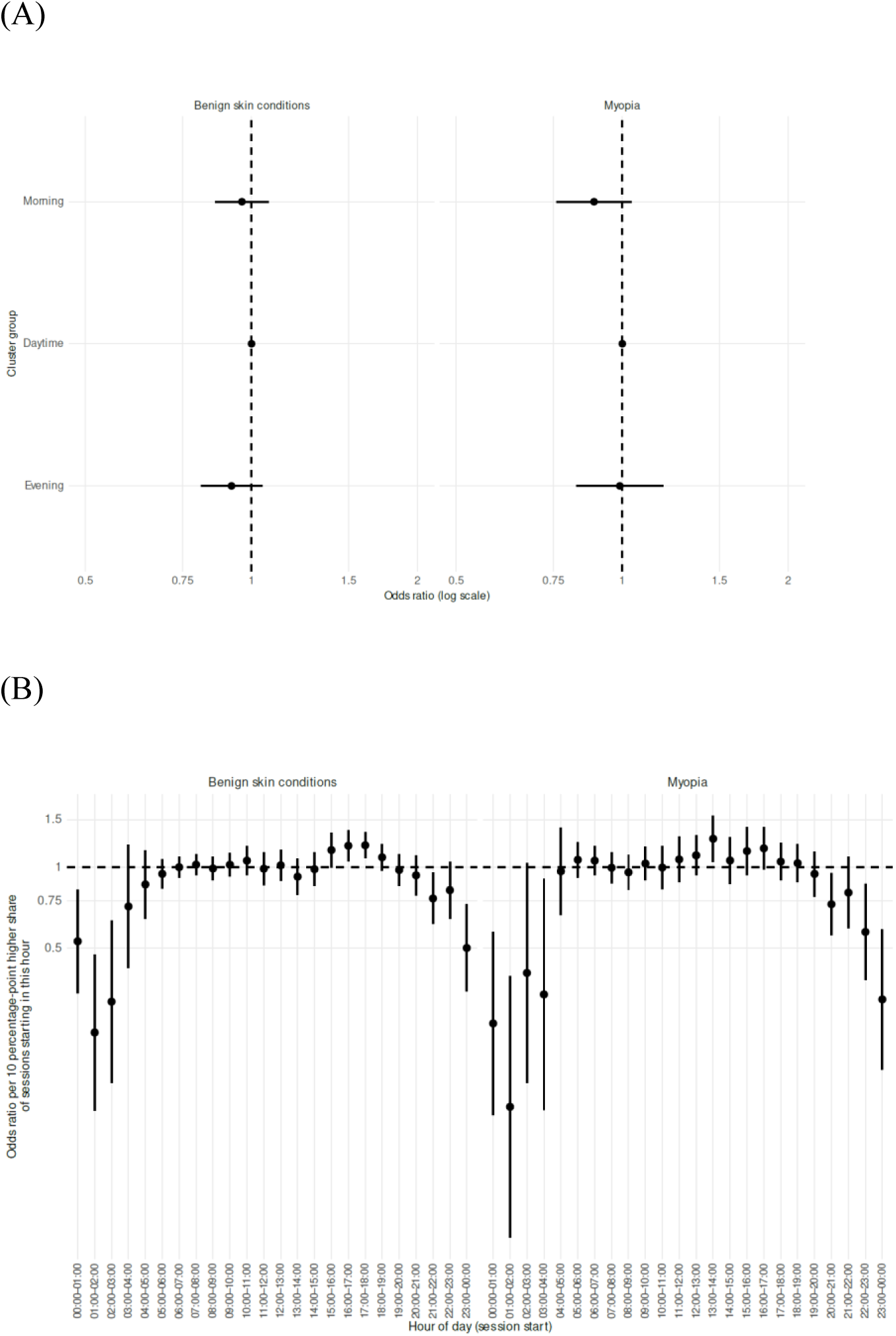
Negative control analyses. Associations between exercise timing phenotypes and non-cardiometabolic outcomes (benign skin conditions, myopia) are shown as negative controls. No significant associations were observed across (A) timing groups or (B) across hour-of-day models, supporting specificity of the main findings.

## Notes

### Competing Interest Statement

The authors have declared no competing interest.

### Author Declarations

The study used ONLY openly available human data that were originally located at https://workbench.researchallofus.org.

## REFERENCES

1. Piercy, K. L. et al. The physical activity guidelines for Americans. Jama 320, 2020–2028 (2018).

2. Fletcher, G. F. et al. Promoting Physical Activity and Exercise JACC Health Promotion Series. Journal of the American College of Cardiology 72, 1622–1639 (2018).

3. Thompson, P. D. et al. Exercise and physical activity in the prevention and treatment of atherosclerotic cardiovascular disease: a statement from the Council on Clinical Cardiology (Subcommittee on Exercise, Rehabilitation, and Prevention) and the Council on Nutrition, Physical. Circulation 107, 3109–3116 (2003).

4. Bellicha, A. et al. Effect of exercise training on weight loss, body composition changes, and weight maintenance in adults with overweight or obesity: An overview of 12 systematic reviews and 149 studies. Obes Rev 22 Suppl 4, e13256 (2021).

5. Piercy, K. L. et al. The Physical Activity Guidelines for Americans. JAMA 320, 2020 (2018).

6. Physical Activity Guidelines for Americans, 2nd edition.

7. Allada, R. & Bass, J. Circadian Mechanisms in Medicine. N Engl J Med 384, 550–561 (2021).

8. Panda, S., Hogenesch, J. B. & Kay, S. A. Circadian rhythms from flies to human. Nature 417, 329–335 (2002).

9. Acosta-Rodríguez, V. A., Rijo-Ferreira, F., Green, C. B. & Takahashi, J. S. Importance of circadian timing for aging and longevity. Nat Commun 12, 2862 (2021).

10. Sato, S. et al. Time of Exercise Specifies the Impact on Muscle Metabolic Pathways and Systemic Energy Homeostasis. Cell Metabolism 30, 92–110.e4 (2019).

11. Ezagouri, S. et al. Physiological and Molecular Dissection of Daily Variance in Exercise Capacity. Cell Metabolism 30, 78–91.e4 (2019).

12. Chaix, A. & Panda, S. Timing tweaks exercise. Nat Rev Endocrinol 15, 440–441 (2019).

13. Li, X. et al. Effects of the Timing of Intense Physical Activity on Hypertension Risk in a General Population: A UK-Biobank Study. Curr Hypertens Rep 26, 81–90 (2024).

14. Tian, C., Bürki, C., Westerman, K. E. & Patel, C. J. Association between timing and consistency of physical activity and type 2 diabetes: a cohort study on participants of the UK Biobank. Diabetologia 66, 2275–2282 (2023).

15. Albalak, G. et al. Setting your clock: associations between timing of objective physical activity and cardiovascular disease risk in the general population. European Journal of Preventive Cardiology 30, 232–240 (2023).

16. Zhang, Q. Accelerometer-measured physical activity timing and risk of incident atrial fibrillation: a UK Biobank cohort study.

17. Ning, Y., Chen, M., Yang, H. & Jia, J. Accelerometer-measured physical activity timing with incident dementia. Alzheimer’s & Dementia 21, e14452 (2025).

18. Feng, H. et al. Associations of timing of physical activity with all-cause and cause-specific mortality in a prospective cohort study. Nat Commun 14, 930 (2023).

19. Leota, J. et al. Dose-response relationship between evening exercise and sleep. Nat Commun 16, 3297 (2025).

20. Master, H. et al. Association of step counts over time with the risk of chronic disease in the All of Us Research Program. Nat Med 28, 2301–2308 (2022).

21. The “All of Us” Research Program. The new england journal of medicine (2019).

22. Chomistek, A. K., Shiroma, E. J. & Lee, I.-M. The Relationship Between Time of Day of Physical Activity and Obesity in Older Women. Journal of Physical Activity and Health 13, 416–418 (2016).

23. Alizadeh, Z., Younespour, S., Rajabian Tabesh, M. & Haghravan, S. Comparison between the effect of 6 weeks of morning or evening aerobic exercise on appetite and anthropometric indices: a randomized controlled trial. Clinical Obesity 7, 157–165 (2017).

24. Blankenship, J. M. et al. Examining the Role of Exercise Timing in Weight Management: A Review. Int J Sports Med 42, 967–978 (2021).

25. Schumacher, L. M., Thomas, J. G., Raynor, H. A., Rhodes, R. E. & Bond, D. S. Consistent Morning Exercise May Be Beneficial for Individuals With Obesity. Exercise and Sport Sciences Reviews 48, 201–208 (2020).

26. Morales-Palomo, F. et al. Efficacy of morning versus afternoon aerobic exercise training on reducing metabolic syndrome components: A randomized controlled trial. The Journal of Physiology 602, 6463–6477 (2024).

27. Pendergrast, L. A. et al. Time of day determines postexercise metabolism in mouse adipose tissue. Proc. Natl. Acad. Sci. U.S.A. 120, e2218510120 (2023).

